# Personalizing deep brain stimulation using advanced imaging sequences

**DOI:** 10.1101/2021.10.04.21264488

**Authors:** Clemens Neudorfer, Daniel Kroneberg, Bassam Al-Fatly, Lukas Goede, Dorothee Kübler, Katharina Faust, Ursula van Rienen, Anna Tietze, Thomas Picht, Todd M. Herrington, Erik H. Middlebrooks, Andrea Kühn, Gerd-Helge Schneider, Andreas Horn

## Abstract

**Objective:** With a growing appreciation for interindividual anatomical variability and patient-specific brain connectivity, advanced imaging sequences offer the opportunity to directly visualize anatomical targets for deep brain stimulation (DBS). The lack of quantitative evidence demonstrating their clinical utility, however, has hindered their broad implementation in clinical practice.

**Methods:** Using fast gray matter acquisition T1 inversion recovery (FGATIR) sequences, the present study identified a thalamic hypointensity that holds promise as a visual marker in DBS. To validate the clinical utility of the identified hypointensity we retrospectively analyzed 65 patients (26 female, mean age: 69.1±12.7 years) who underwent DBS in the treatment of essential tremor. We characterized its neuroanatomical substrates and evaluated the hypointensity’s ability to predict clinical outcome using stimulation volume modeling and voxel-wise mapping. Finally, we determined whether the hypointensity marker could predict symptom improvement on a patient-specific level.

**Results:** Anatomical characterization suggested that the identified hypointensity constituted the terminal part of the dentato-rubro-thalamic tract. Overlap between DBS stimulation volumes and the hypointensity in common space significantly correlated with tremor improvement (R^2^=0.16, p=0.017) and distance to hotspots previously reported in the literature (R^2^=0.49, p=7.9e-4). In contrast, the amount of variance explained by other anatomical atlas structures was reduced. When accounting for interindividual neuroanatomical variability, the predictive power of the hypointensity increased further (R^2^=0.37, p=0.002).

**Interpretation:** Our findings introduce and validate a novel imaging-based marker attainable from FGATIR sequences that has the potential to personalize and inform targeting and programming in DBS for essential tremor.

## Introduction

Today, magnetic resonance imaging (MRI) is commonplace in the field of deep brain stimulation (DBS). A vast majority of DBS centers, however, relies on conventional imaging sequences, such as T1w- and T2w-scans, to inform targeting. These sequences have proven effective in clinical practice for the diagnosis and management of neurological conditions, but suffer from inherent shortcomings that impede the reliable visualization of DBS targets.^1^ This holds especially true for the ventral intermediate (VIM) nucleus of the thalamus, which is not readily visible on MRI. In addition, there is mounting evidence that not stimulation of VIM proper, but the ascending dentato-rubro-thalamic (DRT) tract is primarily associated with symptom improvement in ET.^2–5^ Thus, despite the availability and employment of MRI, surgeons continue to rely on indirect targeting approaches – for VIM but also other basal ganglia structures^6^ – estimating the surgical target in relation to fixed and identifiable landmarks, such as the AC-PC plane. This approach, however, fails to sufficiently account for interindividual neuroanatomical variability. In DBS, the employment of intraoperative microelectrode recordings and awake testing, as well as the ability to shape the electric field during chronic stimulation in the postoperative course allow to partially account for potential inaccuracies. While effective, however, these methods are associated with prolonged operative durations and assume multiple penetrations of deep brain structures, thus increasing the risk of intra- and postoperative complications.^7^ Beyond these limitations, the rapid expansion of surgical targets, indications, and treatment strategies that increasingly favor ‘connectomic’ targeting approaches highlights the need for more sophisticated and reliable visualization tools with the ability to account for interindividual neuroanatomical variability.^8,9^ Personalization of DBS targeting may not necessarily increase the benefit of the approach for top-responding patients, but could lead to greater consistency and a *higher fraction* of top-responding patients.

Direct visualization of deep brain structures and DBS targets has markedly improved in recent years. In addition to increases in magnetic field strength, this is owing to optimized acquisition protocols and postprocessing pipelines that improve gray/white-matter contrast or leverage differences in tissue composition, such as iron content.^1^ The resulting sequences – including susceptibility-based techniques^10^ and inversion recovery sequences^11,12^ – have already been adopted in clinical practice by individual groups; a broad implementation across DBS centers is, however, currently lacking.^1^ Given their relative ease of implementation a potential explanation for the sparse employment of these sequences is the lack of quantitative evidence demonstrating their actual clinical utility.

The aim of the present study is to investigate the clinical utility of advanced imaging sequences for DBS targeting in essential tremor (ET). Specifically, we demonstrate that a visual marker that is appreciable on fast gray matter acquisition T1 inversion recovery (FGATIR) sequences i) can be visualized reliably and reproducibly, ii) can predict symptom improvement in a clinical cohort, and iii) may serve as a surrogate marker for a DBS target that yields optimal tremor suppression. The FGATIR sequence was introduced as part of our clinical imaging acquisition protocol for thalamic DBS at our center with the goal to discern individual thalamic nuclei. However, during planning procedures we frequently noticed an oval hypointense marker in close proximity to the identified target at the ventral border of the human motor thalamus. This clinical finding prompted us to retrospectively investigate the neural substrates underlying the marker and probe its predictive utility.

## SUBJECTS/MATERIALS AND METHODS

### Patient selection

Following institutional research ethics board approval, we retrospectively analyzed a total of 65 patients across three cohorts (Fig 1). The first cohort (*BaseVIM Cohort*) featured a total of 36 patients (13 female, mean age: 74.3 ± 11.9 years) suffering from severe, medically intractable essential tremor (ET). These patients had been well characterized in a prior retrospective trial^13^ and underwent bilateral VIM-DBS at Charité – Universitätsmedizin, Berlin between 2001 and 2017. Since *no FGATIR scans* had been acquired in this cohort owing to a lack of imaging protocols at the time, we only derived 12-month follow-up scores and stimulation settings from this cohort. The second cohort comprised ten patients (BerlinNative Cohort; 5 female, mean age: 63.7, range: 39-74 years) who underwent VIM DBS between May 2019 and April 2020 at our center (n=10, the hypointensity was discovered during surgical planning in these patients) and featured patient-specific FGATIR scans for analysis in *native space*. Crucially, while surgeons were not blinded to the FGATIR scans in this cohort, the marker did not alter the planning procedure or stimulation programming (i.e., did not alter clinical practice), since its clinical relevance was unknown. A third cohort operated at Mayo Clinic, Jacksonville, Florida (FloridaNative Cohort; n=19 unilateral implantations, 8 female, mean age: 69.2 years, range: 52-78 years) served as an independent validation cohort. In this cohort the treating physicians (surgeons and programmers) were entirely naïve to the hypointensity and hypothesis. *Patient-specific* FGATIR scans and 12-month follow-up scores in this cohort were retrospectively analyzed to confirm our exploratory findings.

**Figure 1:**
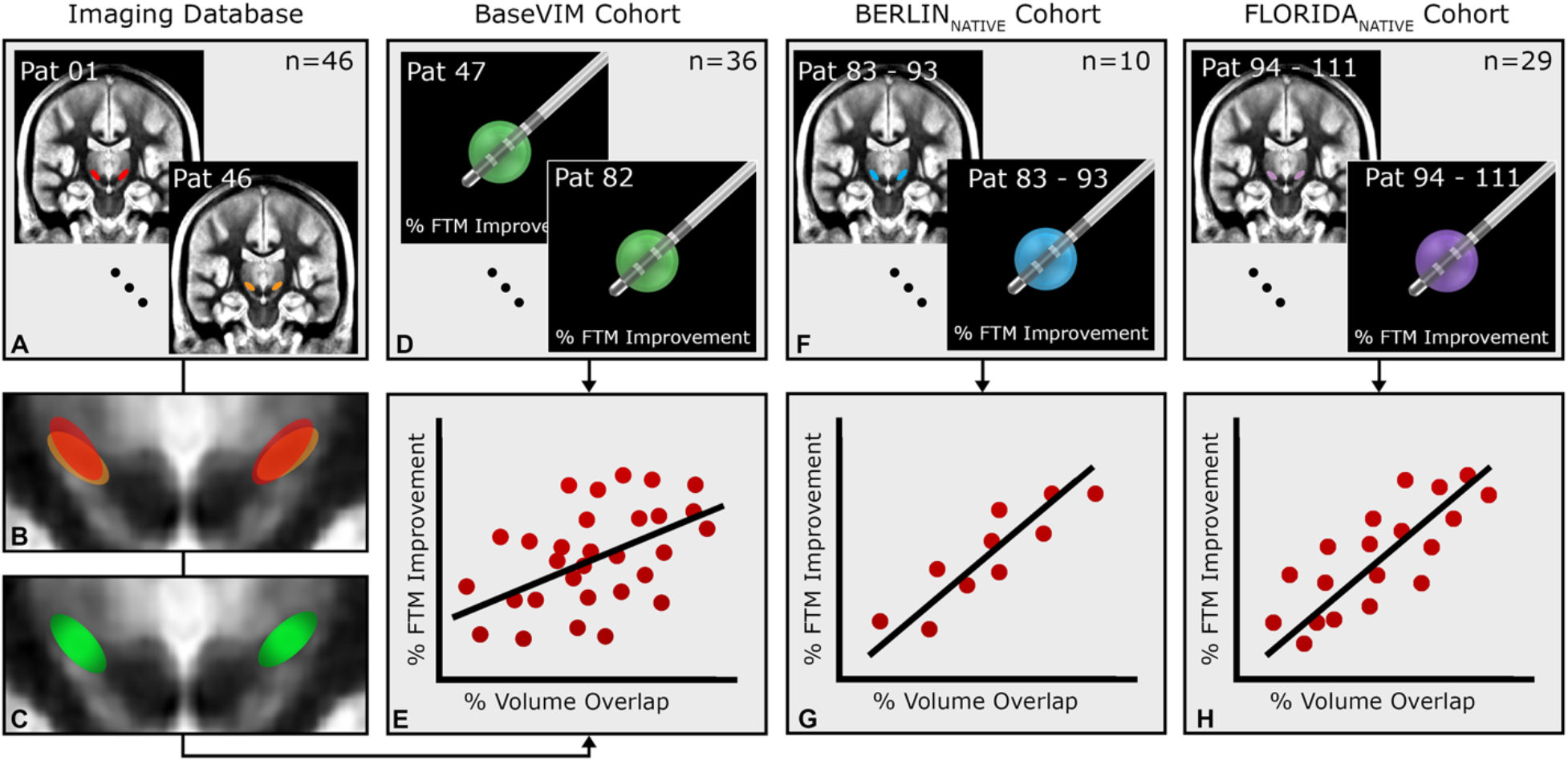
Overview of the cohorts and methods used to investigate the clinical utility of the identified hypointensity. **(A)** We derived FGATIR sequences (n=46) from an imaging database established at Charité – Universitätsmedizin, Berlin to generate a normative template of the hypointensity. To this end hypointensities visible on FGATIR sequences were segmented and transformed into MNI space **(B)**. The hypointensity template was then generated by summing the number of hypointensities overlapping at each voxel and thresholding the resulting n-map at 50% to control for outlier voxels that were only encompassed by a minority of hypointensities **(C)**. Overlap between the final normative hypointensity template and stimulation volumes derived from the *BaseVIM Cohort* (n=36) **(D)** was then calculated and correlated with clinical outcome of the same cohort **(E)**. This approach was taken since no FGATIR sequences had been acquired in the *BaseVIM Cohort*. **(F)** To investigate the predictive ability of the identified hypointensity in patient space we derived two additional cohorts from Berlin and Florida (n=29 patients) where each patient featured both preoperative FGATIR scans, as well as postoperative follow-up scores and stimulation parameters. The volume overlap between each patient’s specific hypointensity and their associated stimulation volume was then correlated with clinical improvement in native space **(G)**. Importantly, this analysis was performed to determine whether the hypointensity would be able to effectively predict clinical outcome taking into account interindividual neuroanatomical variability.

### Imaging Database

Since no FGATIR scans had been acquired in the *BaseVIM Cohort*, we derived preoperative FGATIR sequences from an imaging database established at Charité – Universitätsmedizin, Berlin to correlate clinical outcome in the *BaseVIM Cohort* with hypointensity overlap in common space (Fig 1). In total, we acquired 46 FGATIR scans from our database: 28 scans were obtained from patients undergoing DBS in the treatment of Parkinson’s disease (PD), while 18 scans were obtained from ET patients. The dice similarity coefficient (DSC) was used to compare the spatial overlap of hypointensities across diseases and investigate disease specific differences. The Hausdorff distance was used to measure the maximum Euclidian distance between the closest voxels across corresponding segmentations. Since no clinical data was available in the imaging database only preoperative scans were used in the present study.

### Imaging acquisition

In all patients, high-resolution T1w and T2w MRI scans were obtained preoperatively using a 3.0 Tesla clinical MRI scanner (Skyra Magnetom, Siemens, Erlangen, Germany). In addition, FGATIR sequences were acquired in the *Imaging*, BerlinNative and FloridaNative cohort (n = 75 patients). The complete MRI acquisition protocol consisted of a three-plane localizing scout, a T1-weighted (T1w) 3D Magnetization Prepared-Rapid Acquisition Gradient Echo (MP-RAGE) sequence, a T2w Turbo Spin-Echo (TSE) sequences, and a T1w 3D Fast Gray Matter Acquisition Inversion Recovery (FGATIR) sequence. A detailed overview of the acquisition parameters used for the protocol can be obtained from Table 1. The FGATIR protocol is supplied in the supplementary material.

**Table 1:** Imaging sequences and acquisition parameters employed during preoperative magnetic resonance imaging (MRI). Note that sequences were obtained on a 3.0 Tesla clinical MRI scanner (Skyra Magnetom, Siemens, Erlangen, Germany) and parameters were optimized accordingly. FGATIR can be implemented on 1.5 Tesla scanners, however, parameters would have to be adjusted accordingly.

|  | T1-w 3D MP-RAGE | T2-TSE | T1-w 3D FGATIR |
| --- | --- | --- | --- |
| Repetition time (TR) | 2300 ms | 13320 ms | 3000 ms |
| Echo time (TE) | 2.32 ms | 101 ms | 3.44 ms |
| Inversion time (TI) | 900 ms | n.a. | 414 ms |
| Inversion pulse angle | 90° | n.a. | 180° |
| Field of view (mm) | 240 x 240 | 250 x 250 | 240 x 240 |
| Slices | 192 x 0.9 mm | 70 x 2.0 mm | 160 x 1 mm |
| Orientation | Sagittal | Axial | Axial |
| Bandwidth | 200 Hz/Px | 217 Hz/px | 130 Hz/Px |
| Acquisition time | 5:21 min | 4:15 min | 6:17 min |
| Voxel size | 0.9 x 0.9 x 0.9 | 0.7 x 0.7 x 2.0 | 0.9 x 0.9 x 1.0 |

### Surgical procedure

The surgical techniques associated with VIM-DBS have been reported previously.^13^ In brief, indirect targeting of VIM was performed based on T1w, T2w, and – in the BerlinNative and Florida_Native_ cohort – FGATIR sequences MRI scans. The preliminary target for VIM (reflecting the center of the most distal electrode contact) was identified 13.0-14.0 mm lateral, 6.0 mm anterior, and +/- 0.0 mm ventral to the posterior commissure and adjusted based on available imaging data. Of note, in the BerlinNative cohort the adjusted target frequently coincided with a focal, circumscribed hypointensity on FGATIR sequences, which is subject to the present study (Fig 2). Importantly, while the planning surgeons were not blinded to the FGATIR scan, the hypointensity did not inform targeting as its functional relevance had been unknown prior to this retrospective analysis. Intraoperatively, microelectrode recordings and test stimulation were used to guide lead placement and determine the location of optimal clinical response. To this end, leads were commonly advanced ventrally into the posterior subthalamic area (PSA) to maximize tremor suppression and reduce the currents required for effective symptom control. Following test stimulation, electrodes were internalized and connected to an internal pulse generator under general anesthesia. Computed tomography (CT) imaging was performed postoperatively to clinically validate lead placement with respect to the intraoperatively determined trajectory and surrounding neuroanatomy and to exclude lead displacement.

**Figure 2:**
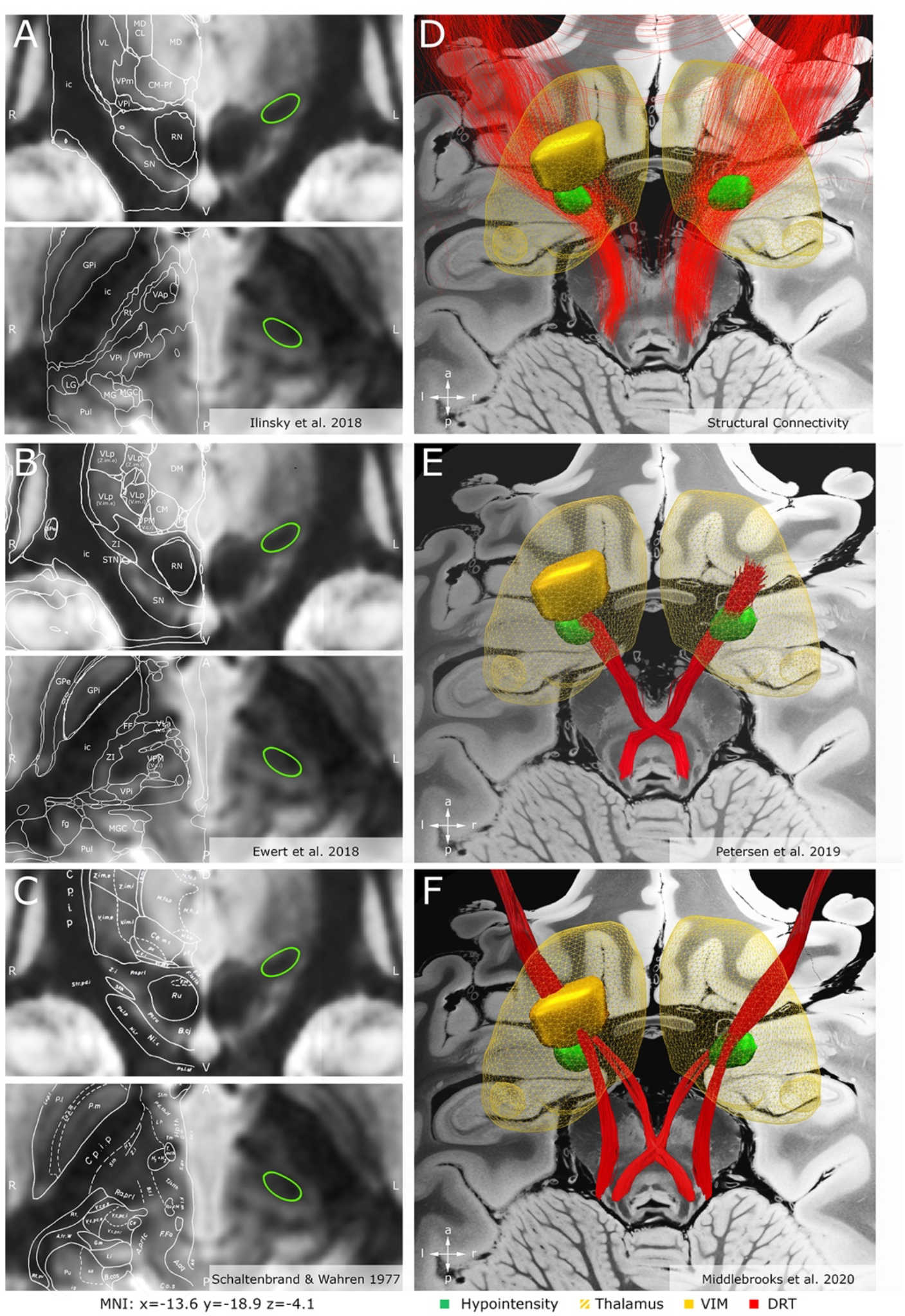
Spatial characterization of the identified thalamic hypointensity in synopsis with histological, stereotactic and fiber tract atlases. **(A-C)** Histological/stereotactic atlases superimposed on coronal (top) and axial (bottom) sections of an average FGATIR template generated from 46 preoperative patient scans (*Imaging Database*) in MNI space. The identified hypointensity (green), which served as a visual marker in the present study featured an oval shape at the level of the posterior subthalamic area (PSA) extending in a posteromedial to anterolateral manner into the ventral aspects of the ventral intermediate (VIM) thalamic nucleus. **(D)** The identified hypointensity was used as a seed region to identify the streamlines traversing the structure. **(E-F)** Superimposition of the visual marker and fiber tract atlases derived from the literature suggests that the dentato-rubro-thalamic (DRT) tract is the likely substrate underlying the hypointensity. Note the convergence of decussating and non-decussating part of DRT within PSA that coincides with the location of the hypointensity.

### Segmentation of hypointensities and template generation

Segmentation of the hypointensity was performed by two raters (CN, LG) who were blinded to clinical outcomes, electrode localizations, and the probabilistic mapping results (see below) and manually labeled the region of interest (ROI) in a total of 75 FGATIR scans. Raters were instructed to segment an oval hypointensity at the base of the thalamus after discussing 2-5 example cases. All labels were generated on unprocessed scans in native space using MNI Display (Montreal Neurological Institute; http://www.bic.mni.mcgill.ca/software/Display/Display.html). Following visual identification of the hypointensity on coronal sections, the ROI was segmented bilaterally in all planes. To determine the extent of inter-rater variability the dice similarity coefficient (DSC) was calculated for all labels following segmentation.

A probabilistic template of the FGATIR hypointensity was generated by transforming native segmentations into ICBM 2009b NLIN Asym (“MNI”) space using Lead-DBS v2.5 (https://www.lead-dbs.org).^14^ To this end multimodal (T1w, T2w, and FGATIR) sequences of each patient were first rigidly co-registered using SPM12 (https://www.fil.ion.ucl.ac.uk/spm/software/spm12) and then non-linearly transformed to MNI space using the ‘effective: low variance + subcortical refinement’ preset for Advanced Normalization Tools (ANTs; http://stnava.github.io/ANTs/). Each normalization was visually validated and, if necessary, refined using an additional subcortical transformation step.^15^ The final transformation matrix of each patient was then used to transform patient-specific segmentations into MNI space. Once transformed, frequency maps reflecting the number of individual segmentations at each voxel were generated. To control for outlier voxels that were only encompassed by a minority of ROIs and to increase the confidence of overlap across ROIs the frequency map was thresholded at 50%.

### Voxel-wise statistical analysis (probabilistic mapping)

To investigate the spatial relationship between our final hypointensity template and the location of best overall tremor improvement we generated probabilistic maps of stimulation volumes in MNI space. The underlying stimulation parameters were acquired retrospectively at 12-month follow-up. Programmers were blinded to the hypointensity and adjustment of stimulation settings was performed empirically according to our center’s standard of care. For analysis, DBS electrodes were first localized in all patients using default parameters of the Lead-DBS pipeline.^14^ Stimulation volumes were then estimated using the SimBio/Fieldtrip^16^ approach as implemented in Lead-DBS using the stimulation settings obtained from each patient at 12-month follow-up. Our probabilistic mapping approach followed the strategy employed by Elias et al.^17^ Briefly, patient-specific stimulation volumes were first weighted by their corresponding relative clinical change from baseline as assessed by the Fahn-Tolosa-Marin (FTM) tremor rating scale. To this end, each voxel included in a patient’s stimulation volume was assigned the relative improvement score observed in this patient (e.g. the value 0.8 was assigned to the voxel in case of 80% tremor improvement). These values were then aggregated across patients in a voxel-wise fashion after i) demeaning and dividing the values by the volume (in cubic mm) of the stimulation volume to penalize larger, less focal stimulation volumes. For each voxel, the group mean was then computed by averaging the sum of all weighted volumes overlapping a given voxel – this process produced a raw average map. To control for outlier voxels that were only encompassed by a minority of volumes, an unweighted *n-*map was generated that featured the total number of volumes overlapping each voxel. The *n*-map was conservatively thresholded at 10% and then used to mask the raw average map. Finally, voxel-wise, two-tailed Wilcoxon signed-rank tests were performed to calculate a *p*-map. This map indicated the degree of confidence for electrical stimulation at each voxel to be associated with clinical change with respect to the cohort-specific average improvement. The final *p*-map was thresholded at p<0.05 to only retain voxels significantly associated with clinical change and used to mask the *n*-masked average map.

### Dataset validation

To determine whether our hypointensity template could be used as a predictor for clinical outcome in surgical planning and postoperative programming we calculated the overlap between each stimulation volume and the hypointensity template in MNI space. We then generated linear models using volume overlap as dependent variable to explain variance in clinical outcome (total tremor improvement). This process was repeated for alternative diencephalic structures including VIM^18^, DRT^2^, as well as the previously generated probabilistic maps to compare overall model performance. Because patients were stimulated bilaterally, the percent overlap between ROIs and stimulation volumes was aggregated across both hemispheres in each patient. In a separate analysis, we derived hotspots and target coordinates from previous studies investigating the efficacy of VIM-DBS in ET. Both retrospective and prospective studies were included if they I) reported target coordinates or hotspots in MNI space or with respect to AC-PC, II) provided average clinical outcome scores associated with tremor suppression in the postoperative course, and III) featured cohorts comprising at least ten patients. AC-PC coordinates were converted into MNI coordinates as previously described.^19^ We then calculated the average Euclidean distance from the reported peak intensities/target coordinates to the centers of gravity of our hypointensity template in both hemispheres. Finally, linear regression was performed to assess the relationship between reported total improvement scores and proximity to the hypointensity.

### Patient-specific validation in native and MNI space

To make statistical inferences on *a group level* in neuroimaging the transformation of native patient data into common space is required. However, this process is associated with an inherent loss in precision and disregards interindividual neuroanatomical variability. Hence, a spatial marker that could be reliably detected in each specific patient (without the need of atlas registrations) would be of great value. Unfortunately, no preoperative FGATIR scans were available in the *BaseVIM* cohort, so two additional, independent cohorts (BerlinNative and FloridaNative) were analyzed that featured both preoperative FGATIR sequences and clinical outcomes after one year of continuous stimulation (Fig 1). Here, overlaps between each patient’s individual stimulation volume and their respective hypointensity marker were correlated with clinical outcome in native space without any nonlinear registration steps involved. This process was then repeated in MNI space following normalization of patient-specific segmentations to compare changes in predictive power. To determine whether correlations were non-random, non-parametric permutation analysis was performed.

## RESULTS

### Anatomical characterization of the identified hypointensity

During surgical trajectory planning the final target frequently coincided with a hypointense oval region on FGATIR sequences extending from the level of AC-PC ventrally into the posterior subthalamic area (PSA). The hypointensity was consistently and reproducibly identifiable (mean interrater DSC: 0.8 ± 0.04, mean Hausdorff distance: 1.7 ± 0.48 mm). On a group level, its bilateral centers of gravity were located at MNI coordinates x = 12.57, y = -17.10, z = -3.45 mm and x = - 13.24, y = -18.34, z = -3.23 mm, respectively, with an average volume of 124.9 ± 35.2 mm^3^. Comparing the spatial overlap of segmentations across diseases (PD imaging dataset vs ET imaging dataset) revealed a DSC of 0.70 suggesting a uniform topology of identified hypointensities across conditions.

Traversing PSA in anterolateral to posteromedial direction, the hypointensity adjoined the posterior border of the subthalamic nucleus (STN) and zona incerta (ZI) rostrally, while – based on aggregated information of several anatomical atlases^20–22^ – being caudally confined by the ventral aspects of ventral posteromedial (VPM) and ventral posterior inferior (VPI) thalamic nuclei, partially extending into the thalamic gray matter (Fig 2A-C). While the hypointensity was clearly separable from ZI, its ventromedial aspect frequently intersected with the hypointensity of the RN, at times becoming indistinguishable from the latter (Fig 2). This description is consistent with the anatomical characterization of the passage of DRT within the PSA.^23,24^ To confirm that the identified hypointensity indeed overlapped with the terminal part of DRT, we superimposed the segmented hypointensity template on established histological^20,21^, stereotactic^22^, and white matter atlases^25,26^ (Fig 2). Furthermore, the template was used as a seed region to identify streamlines passing through the hypointensity (Fig 2D). Taken together, this suggested that the identified hypointensity could indeed constitute the most terminal aspect of DRT.^12^ Of note, Schaltenbrand and Wahren (Fig 2C) associated the identified hypointensity with the prelemniscal radiation (Raprl), a white matter region within the PSA comprising i) DRT and its associated projections to primary and supplementary motor cortex, ii) fibers ascending from the mesencephalic reticular formation to the orbitofrontal and prefrontal cortices, and potentially iii) streamlines connecting the pedunculopontine nucleus and the pallidum.^23,27^ Within Raprl, however, the anti-tremor effects are most likely associated with DRT stimulation.

### Probabilistic stimulation mapping

To determine whether the identified hypointensity could serve as a useful spatial marker for surgical planning and DBS programming, its ability to predict symptom improvement following DBS was evaluated. In a first, step, we created optimal treatment targets by applying a probabilistic mapping approach^17^ to the *BaseVIM* (N = 36) cohort. Inspection of these maps revealed spatially distinct clusters at the level of the ventral thalamus and PSA that were associated with above-mean tremor improvement (≥65.1%) and precisely matched the location and extent of the hypointensity template in MNI space (‘hotspot’, Fig 3D-F). In total, 46.9% of positively correlating voxels overlapped with the hypointensity, while the remaining predictive voxels adjoined the segmentation anterodorsally extending into the ventral lateral posterior (VLpv) thalamic nucleus. The peak intensities of both left- and right-sided hotspots were fully encompassed by their respective hypointensity templates (Fig 3D-F). By contrast, both a more anterior (left) region of the subthalamic area and a circumscribed area ventral to the ROI (right) were associated with below-mean overall clinical improvement (‘coldspot’, Fig 3D-F). These voxels did not touch the hypointensity and extended into ZI.

**Figure 3:**
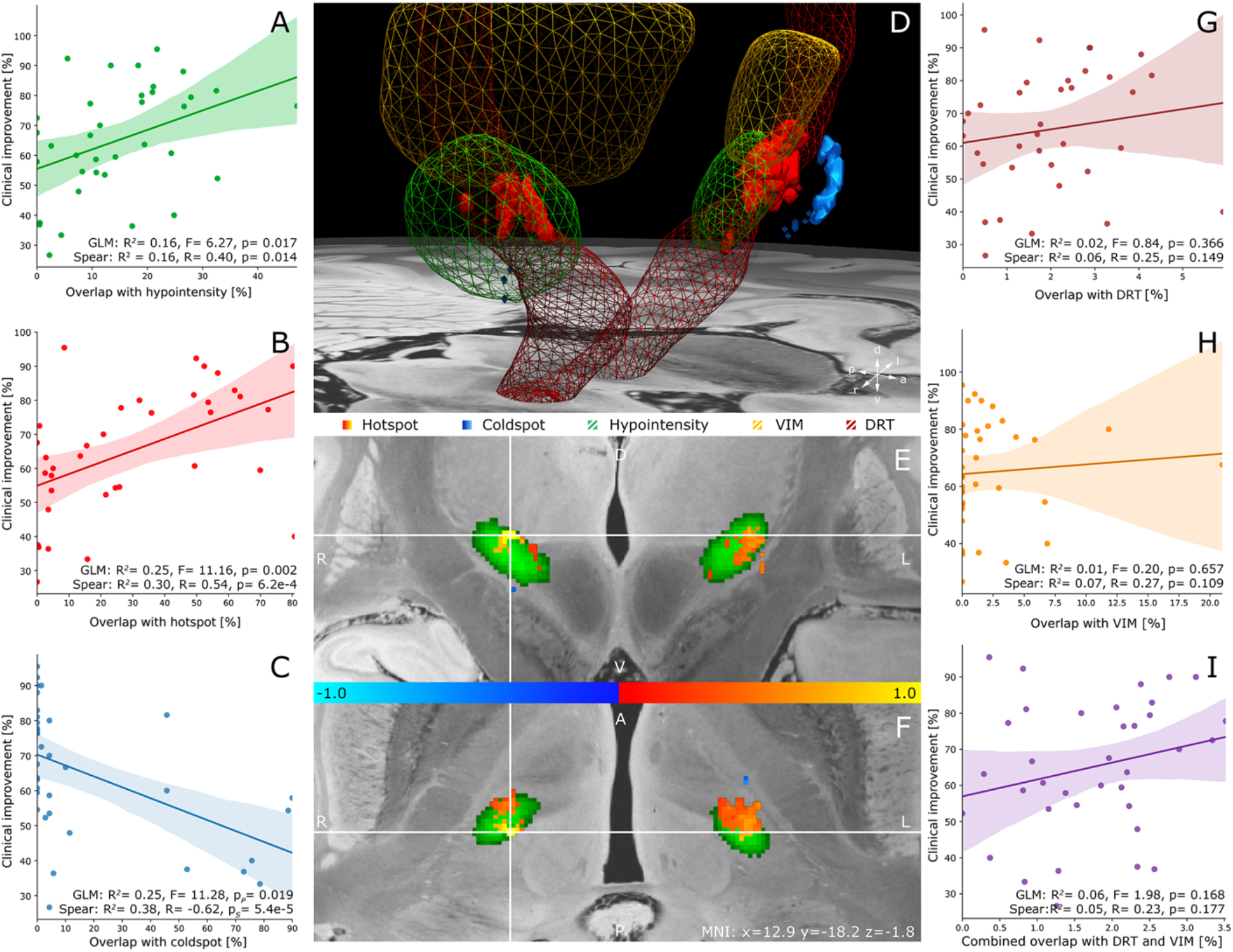
Probabilistic stimulation maps identifying brain areas associated with effective tremor suppression and validation. Clusters that were significantly associated with above-mean (hotspot, red) and bellow-mean (coldspot, blue) tremor improvement overlaid with the identified thalamic hypointensity (green) are projected on axial **(D, F)** and coronal **(E)** slices of a 100-micron resolution, 7T brain scan in MNI 152 space. The slices in E and F are centered on the right-sided peak intensity (MNI: x=12.9 y=-18.2 z=-1.8), which coincided with the dorsal aspect of the hypointensity. **(A-C)** Validation of probabilistic stimulation maps and hypointensity template in the *BaseVIM cohort*. For each structure the relationship between clinical improvement and the extent of stimulation volume overlap is shown: (A) hypointensity, (B) hotspot, (C) coldspot. **(G-I)** Validation of atlas structures including the dentato-rubro-thalamic tract, ventral intermediate (VIM) thalamic nucleus, and both structures combined (i.e., cerebellothalamic outflow tract) in the *BaseVIM cohort*. While stimulation volume overlap with the hypointensity could explain a significant amount of variance in clinical improvement, no such relationship could be established for other atlas structures, emphasizing the clinical importance of this area in effective tremor suppression. (G) DRT, (H) VIM, (I) cerebellothalamic outflow tract (DRT+VIM). GLM: General linear model, Spear: Spearman’s Correlation Coefficient.

### Validation of probabilistic mapping

We next investigated the predictive power of the hypointensity with respect to clinical improvement and compared the model’s performance to correlations with hotspot, coldspot, VIM, DRT, and other structures which have been implicated in tremor suppression, namely pallidothalamic tract^28^ and caudal zona incerta (cZI).^29,30^ To this end, simple linear models were generated investigating the relationship between percent volume overlap and clinical improvement scores obtained at 12 months compared to preoperative baseline (Fig 3). Overlap between each patient’s stimulation volume and the hypointensity template significantly correlated with clinical improvement (R^2^=0.16, p=0.017). Correlation based on the hypointensity template generated from segmentations by the *second tracer* (test-retest reliability) revealed similar results (R^2^=0.13, p=0.029).

Conversely, models using anatomical atlas structures, namely VIM-, DRT-, and a combination of both structures (i.e., cerebellothalamic outflow tract) did not demonstrate a significant relationship with outcomes (VIM: R^2^=0.01, p=0.657; DRT: R^2^=0.02, p=0.366; Cerebellothalamic outflow tract: R^2^=0.06, p=0.168; Fig 3G-I). A similar relationship was observed with respect to pallidothalamic tract (R^2^=0.29, R=-0.54, p=7.6e-4) and cZI (R^2^=0.02, p=0.471). The amount of variance explained by overlap with the identified hot- and coldspots was greatest featuring a positive correlation with hotspot overlap (R^2^=0.25, p=0.002) and a negative correlation with coldspot overlap (R^2^=0.25, p=0.019). Crucially, this latter analysis has to be considered circular given that hot- and cold-spots were defined and validated on the same – *BaseVIM* – cohort of patients. In contrast, the hypointensity template was derived from an independent dataset (Imaging database), suggesting a clinically meaningful relationship between hypointensity overlap and clinical outcome.

### Comparison to other hotspots

To further characterize the predictive potential of the hypointensity marker, we determined its spatial relationship to previously published hotspots in the literature (Table 2). The identified coordinates closely followed the trajectory of the hypointensity template demonstrating an anterolateral to posteromedial course (Fig 4). As a result, ventral stimulations primarily extended into PSA, just inferior to VIM, while dorsal stimulations were encompassed by the thalamus extending into VLp. With the exception of the peak intensity identified by Elias et al.^17^, which was located at the anterior border of VIM and encroached on VLp, all coordinates overlapped with or were in close proximity to the hypointensity template (Fig 4). The mean Euclidean distance from each coordinate to the center of gravity of the hypointensity was 3.8 ± 2.2 mm. We found a strong negative correlation between reported clinical outcomes in respective studies and the distance from their identified hotspots to the hypointensity’s center of gravity (R^2^=0.49, p=7.9e-4; Fig 4C). In other words, the closer a reported hotspot was to the hypointensity, the greater the reported tremor improvement was.

**Table 2:** Overview of previously published hotspots/targets in the literature. To investigate the spatial relationship of our identified hypointensity to previously identified targets we performed a literature search including studies that reported both target coordinates and clinical outcomes in ET patients undergoing Vim DBS. Coordinates reported with respect to AC-PC were converted to MNI space as previously described.^38^ ETRS essential tremor rating scale; FTM, Fahn-Tolosa-Marin tremor rating scale; MNI, Montreal Neurological Institute.

| Reference | Study Type | No. of patients | Follow-up period [Mo] | Coordinates (x/y/z) [mm] | Space | Outcome measure | Improvement [%] |
| --- | --- | --- | --- | --- | --- | --- | --- |
| Papavassiliou et al.<sup>31</sup> | Retrospective | 37 | 26 | -14.5/-17.7/-2.8 | MNI | FTM (limited) | 53.0 |
| Hamel et al.<sup>32</sup> | Retrospective | 10 | 12 | -12.7/-7.0/-1.5 | AC-PC | FTM (total) | 70.1 |
| Herzog et al.<sup>33</sup> | Prospective | 10 | 18.4 | -13.0/-5.5/0.0 | AC-PC | Kinematic analysis | 64.2 |
| Blomstedt et al.<sup>4</sup> | Retrospective | 21 | 12 | -11.6/-6.3/-2.3 | AC-PC | ETRS (total) | 60.0 |
| Barbe et al.<sup>34</sup> | Retrospective | 21 | 3 | -11.3/-7.2/-1.4 | AC-PC | Kinematic analysis | 65.0 |
| Sandvik et al.<sup>24</sup> | Retrospective | 17 | 66 | -13.0/-1.8/4.1 | AC-PC | ETRS (total) | 48.4 |
| Sandvik et al.<sup>24</sup> | Retrospective | 19 | 12 | -12.1/-5.5/-1.2 | AC-PC | ETRS (total) | 58.2 |
| Fytagoridis et al.<sup>29</sup> | Prospective | 50 | 12 | -11.9/-6.2/-2.0 | AC-PC | ETRS (total) | 59.5 |
| Cury et al.<sup>35</sup> | Retrospective | 38 | 12 | -14.7/7.1/1.8 | AC-PC | FTM (total) | 66.0 |
| Fiechter et al.<sup>36</sup> | Retrospective | 12 | 5-22 | -14.3/-5.0/0.9 | AC-PC | FTM (total) | 54.0 |
| Barbe et al.<sup>37</sup> | Prospective | 13 | 12 | -11.0/-5.7/-1.9 | AC-PC | FTM (total) | 64.0 |
| Nowacki et al.<sup>5</sup> | Retrospective | 21 | 12 | -10.6/-5.2/-3.2 | AC-PC | FTM (total) | 55.0 |
| Nowacki et al.<sup>38</sup> | Retrospective | 15 | 12 | -12.8/-3.6/0.0 | AC-PC | Bain and Findley Score | 63.0 |
| Philipson et al.<sup>30</sup> | Retrospective | 26 | 12 | -12.0/-7.5/-4.0 | AC-PC | ETRS (total) | 63.0 |
| Tsuboi et al.<sup>39</sup> | Retrospective | 97 | 12 | -14.3/-4.3/-2.1 | AC-PC | FTM (total) | 53.2 |
| Elias et al.<sup>17</sup> | Retrospective | 39 | 16.8 | -17.3/-13.9/ 4.2 | MNI | FTM (total) | 42.8 |
| Tsuboi et al.<sup>28</sup> | Retrospective | 20 | 6.6 | -15.0/-17.0/ 1.0 | MNI | FTM (total) | 38.7 |
| Middlebrooks et al.<sup>40</sup> | Retrospective | 84 | 6.8 | -15.5/-15.5/ 0.5 | MNI | FTM (total) | 54.6 |

**Figure 4:**
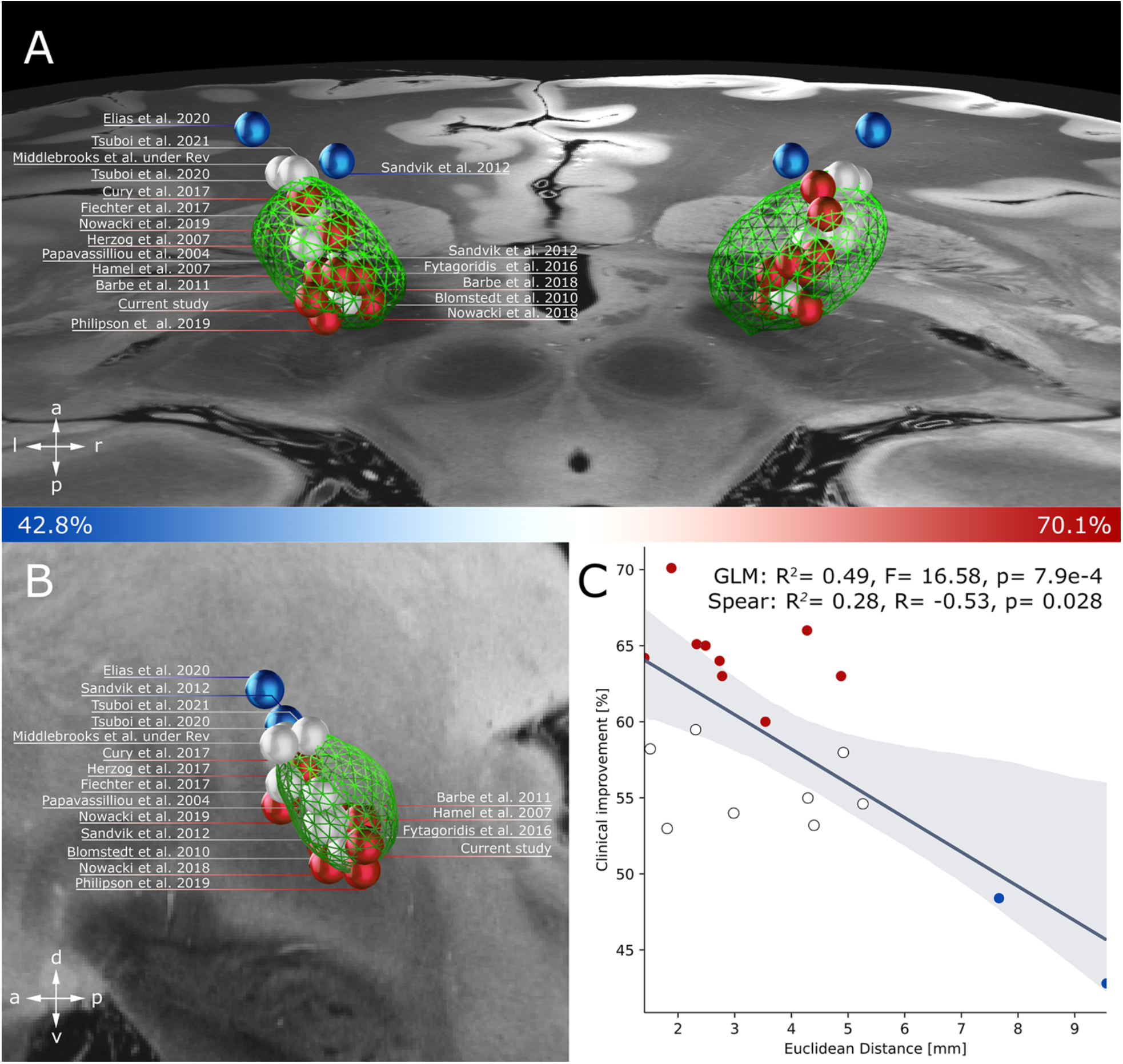
Spatial relationship between the identified hypointensity and targets previously reported in the literature. **(A-B)** The hypointensity (green) is featured on axial and sagittal sections of a 100-micron resolution, 7T brain scan in MNI 152 space. Previously identified hotspots investigating tremor suppression in ET patients following DBS were transformed into MNI space and color-code based on the reported clinical improvement. Targets associated with improved clinical outcome featured a close relationship to the hypointensity frequently coinciding with the structure. For an overview of study characteristics, MNI coordinates, and clinical outcome refer to Table 2. **(C)** Linear regression identified a significant relationship between clinical outcome and the Euclidean distance between target and hypointensity. Overall greater improvement occurred in close proximity to the center of gravity of our visual marker. GLM: General linear model, Spear: Spearman’s Correlation Coefficient.

### Utility of the hypointensity on a patient-specific level

Both probabilistic stimulation mapping and comparison to other hotspots established the predictive utility of the identified hypointensity with respect to tremor improvement. However, given that up to this point all inferences were made on a group level in (normative) MNI space these findings do not address the question of how reliable and useful the hypointensity marker could be when used in individual patients.

To address this question, we aggregated a subset of patients from two different centers (Charité - Universitätsmedizin Berlin and Mayo Clinic, Jacksonville, Florida) who i) had undergone VIM-DBS, ii) had received FGATIR imaging in the preoperative course, and iii) had a follow-up period of 12 months with documented stimulation parameters and outcome measures. Aggregating these patients allowed us to overlap each patient’s individual hypointensity with their respective DBS stimulation volumes in *native* space accounting for the full extent of each patient’s unique anatomical features. Importantly, different planning and stimulation programming strategies were employed across centers, thus allowing a better generalization of the hypointensity’s overall clinical utility.

Exceeding the predictive power of the hypointensity in common space (Fig. 3), this fully individualized model demonstrated a strong association both in the Berlin (R^2^=0.53, p=8.7e-4) and the Florida (R^2^=0.49, p=8.7e-4) cohort, respectively (Fig. 5). Non-parametric permutation analysis confirmed a significant, non-random relationship between volume overlap and clinical outcome in both cohorts (Berlin: p_Permutation_=0.001; Florida: p_Permutation_ < 0.001, n=1000 permutations). To validate this finding, we performed cross-validation, training a linear model on the Berlin cohort and cross-predicting outcome in the Florida cohort (R^2^=0.58, p=1.6e-3). Cross-prediction of the Florida cohort based on the Berlin cohort yielded a similar correlation strength (R^2^=0.53, p=0.016) between reported and predicted outcomes.

**Figure 5:**
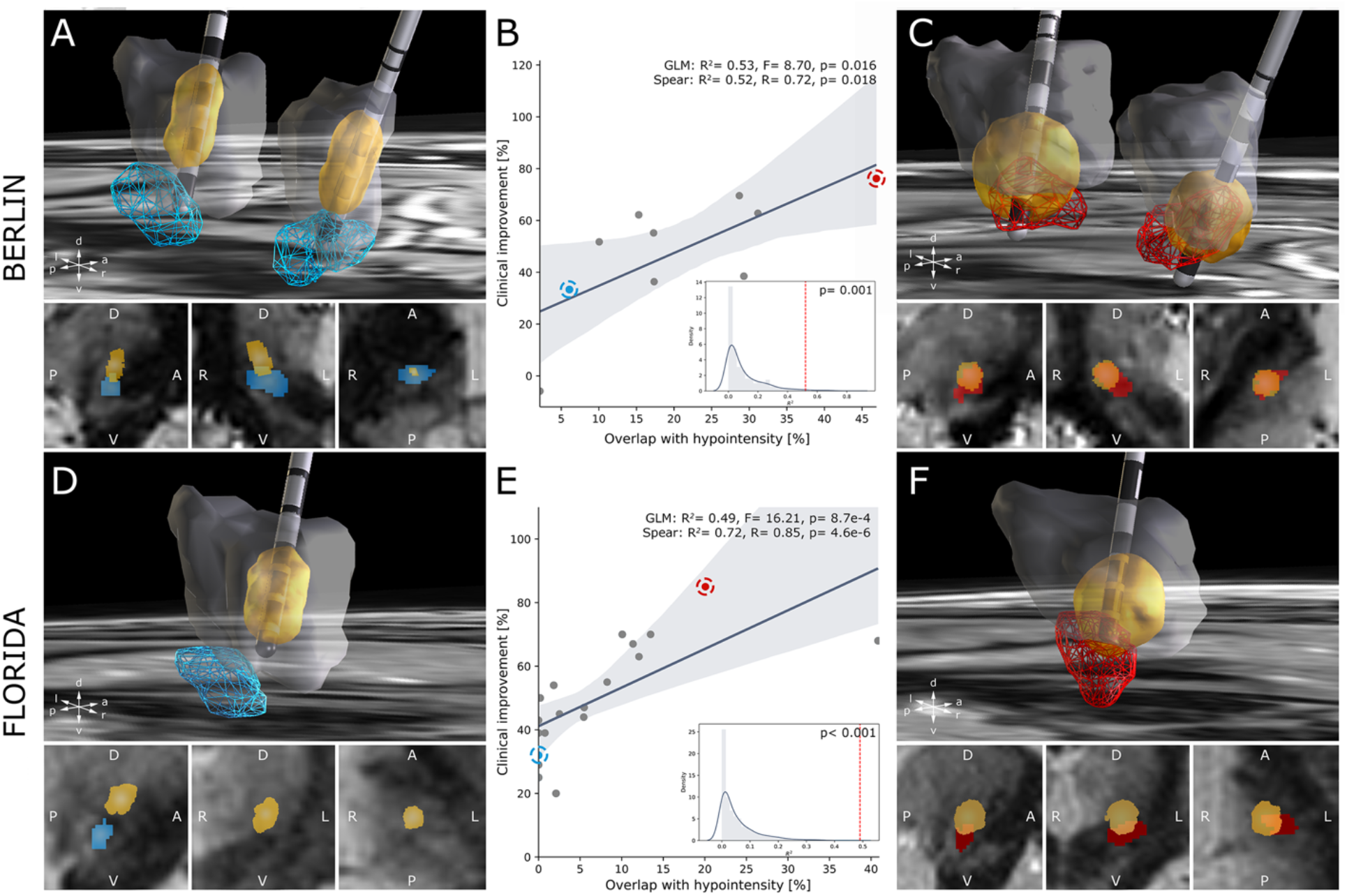
Patient-specific validation of the identified hypointensity in native space. Taking into account interindividual neuroanatomical variability, volume overlap between patient-specific hypointensities and associated stimulation volumes was correlated with clinical outcome in native space across two cohorts: Berlin (**A-C**) and Florida (**D-F**). Note that implantations in the Florida cohort were performed unilaterally; correlations in the figure are reported with respect to the patients’ total FTM improvement scores; analysis of hemi-scores in the Florida cohort revealed similar correlation strengths in native space (GLM: R^2^= 0.38, p = 5.2e-3, p_Permutation_< 0.001, n = 1000 permutations; Spearman’s correlation: R = 0.80, p = 2.9e-5, p_Permutation_<0.001, n = 1000 permutations).Spatial relationships in example poor responding **(A, D)** and top responding **(C, F)** patients are superimposed on the respective patients’ FGATIR sequences in the sagittal (bottom left), coronal (bottom middle), and axial (bottom right) plane. GLM: General linear model, Spear: Spearman’s Correlation Coefficient.

A combined model across all 29 patients was able to explain 51% of the observed variance in clinical outcome (R^2^=0.51, p=1.0e-4, p_Permutation_<0.001, n=1000 permutations) when controlling for cohort (Fig. 6). Correlation based on patient-specific segmentations by the *second tracer* (test-retest reliability) was consistent with this finding (R^2^=0.37, p=0.003, p_Permutation_<0.001, n=1000 permutations). Furthermore, comparison of outcomes between patients whose stimulation volumes overlapped vs. did not overlap with the hypointensity revealed a significant difference (p=0.007, two-sample t-test). Thus, a significantly better outcome can be expected when electrical stimulation encompasses the hypointensity marker. A significant relationship was maintained when warping stimulation volumes and patient-specific hypointensities into MNI space (GLM: R^2^=0.52, p=7.5e-5, p_Permutation_<0.001, n=1000 permutations) (Fig. 6). Overall, these findings suggest that interindividual differences in patient anatomy have a significant influence on the extent of variance explained in outcome. While transformation of individualized models into MNI space is able to maintain this relationship yielding similar correlation strength (due to preservation of interindividual differences), the employment of group-level atlases (Fig. 3) can only account for a fraction of the variance observed, while at the same time requiring larger patient numbers to reach statistical power.

**Figure 6:**
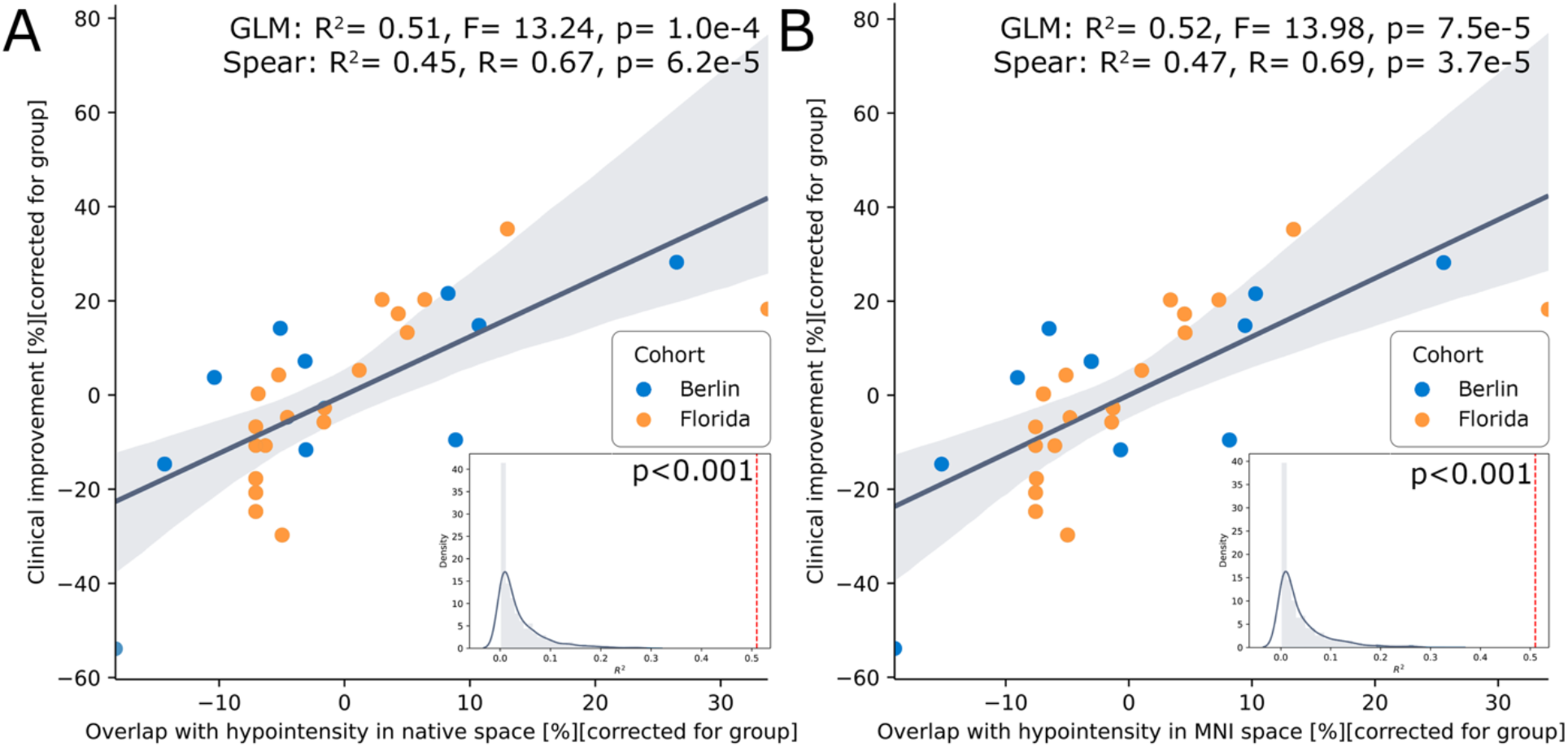
Correlation plots comparing model performance in common (MNI) space and native space. Combined models comprising a total of 29 patients from Berlin (n=10) and Florida (n=19) feature the relationship between clinical outcome and volume overlap in native space **(A)** and following transformation of patient-specific hypointensities and VTAs into MNI space **(B)**. Note that due to preservation of interindividual differences a significant relationship is maintained during normalization, i.e., the exact same data is analyzed but after it was warped to a different space (B). This should not be confused with overlaps between individual VTAs and atlases/group-level templates (see Figure 3). GLM: General linear model, Spear: Spearman’s Correlation Coefficient.

## DISCUSSION

Using advanced imaging sequences, the present study identified an anatomical structure at the level of the posterior subthalamic area encroaching on VIM that may serve as a surrogate marker for optimal tremor suppression during DBS targeting. Specifically, we found that an oval shaped hypointensity at the level of the ventral thalamus could be reliably identified on FGATIR sequences and was able explain a significant amount of variance in observed clinical outcome following VIM-DBS. The marker featured a strong spatial proximity to previously identified hotspots in the literature and could explain differences in clinical effects across studies. Importantly, the predictive power of the hypointensity was greatest when accounting for interindividual neuroanatomical variability in each single DBS patient, rather than using an aggregated template of the marker defined on a group level (Fig 5). Our findings mark an important step beyond experiential evidence in employing an advanced imaging sequence to clinical DBS datasets, providing quantitative evidence of their clinical utility and ability to personalize targeting.

The “oscillating network hypothesis” implicates distributed central oscillators in the pathophysiology of ET that reside within cerebello-thalamo-cortical nodes and synchronize to a tremor-specific frequency.^41^ While electrical disruption of pathological activity and alleviation of symptoms can be assumed within any node of this network, the optimal DBS target would likely be located within a central hub with extensive connectivity to upstream and downstream structures. VIM effectively meets these requirements, its large volume, however, impedes a uniform and effective stimulation of the nucleus proper. In the present study, this might be reflected in the reduced predictive ability of VIM with respect to outcome. This finding is in agreement with an accumulating body of evidence suggesting that not VIM, but specific areas within the posterior subthalamic area may be the most suitable target for effective tremor control.^2,13,32,33,37^ Specifically, ascending cerebellothalamic fibers are confined to a narrow space within DRT at this level forming a bottleneck that can be targeted and modulated effectively.

While we are unable to determine the neuroanatomical structure underlying the identified hypointensity marker with certainty, the synopsis of FGATIR sequences with both fiber-tract and anatomical atlases indicates that DRT proper could be the likely anatomical substrate. Overlap between DBS stimulation volumes and the hypointensity effectively predicted significant amounts of variance in clinical outcome across cohorts, emphasizing its clinical value as a visual marker. Furthermore, under the assumption that the identified hypointensity constitutes DRT, this would underline the notion that electrical stimulation of the cerebellothalamic outflow tract at the level of PSA, specifically within the spatial constraints of the visual marker, is associated with the most distinct tremor suppression.^3,13^ The reduced degree of variance explained by overlap with *atlases* of the DRT^2,26^, however contradicts this assumption. A potential explanation for this finding could be that not all levels of DRT yield equal symptom suppression. Indeed, proximity of DRT to the internal capsule, medial lemniscus, and red nucleus at more ventral and posterior levels could make DBS more prone to off-target side effects, such as ataxia, dysarthria, dysmetria, and paresthesia.^37^ The occurrence of these adverse events, in return, has the potential to drastically reduce the therapeutic window of stimulation at lower levels and might explain the restriction of greatest improvement to the confines of our hypointensity. Furthermore, the hypointensity could also be an expression of locally altered bioelectrical properties that might have direct effects on stimulation efficacy. The notion that the hypointensity not only allowed us to visualize DRT, but also guided us towards its most effective subpart could represent a serendipitous coincidence. Crucially, however, while retrospective screening of patient records in the Berlin and Florida cohorts revealed no intolerable side effects at 12-months follow-up, the lack of recorded minor stimulation-induced adverse events prevented their direct investigation in the present study (similar to several recent studies suggesting efficient tremor suppression along the entirety of DRT).^2,42^ Further prospective validation is required to characterize the side-effect profile of the hypointensity marker.

Recognizing the potential of DRT stimulation and based on a growing appreciation that optimal targets in DBS may feature interindividual neuroanatomical variability, significant effort has been directed toward directly visualizing the cerebello-thalamo-cortical network in recent years.^12,43^ Diffusion MRI (dMRI) in particular has emerged as a powerful tool to visualize DRT, inform patient-specific targeting, and evaluate surgical outcome.^3,13,43^ Diffusion-based imaging, however, is not without limitations. For example, in regions where fibers cross, branch, kiss, splay, or twist tensor models perform poorly and artefactual reconstructions of pathways with false positives and false negatives are likely to occur (see also Fig 2D). In addition, most diffusion sequences apply echo-planar imaging concepts which suffer from stark distortion artifacts that are most pronounced in central parts of the brain. These limitations have instigated the development of alternative approaches such as spherical deconvolution and probabilistic diffusion tractography with multiple fiber orientations.^44^ The implementation of these methods, however, requires dedicated hardware, scanning time, and post-processing pipelines – requirements that exceed the resources of most clinical centers. These methodological limitations coupled with a low test-retest reliability of tractography results might also explain the lack of prospective trials investigating the clinical utility of tractography. Indeed, dMRI-based targeting has thus far only been demonstrated in small case series. Thus, while dMRI provides the opportunity to uncover unique information about structural connections within the brain and constitutes an integral part in neuroimaging research, current limitations restrict its broad employment in clinical practice.

Advanced imaging sequences – such as FGATIR or related sequences^1^ – could overcome several limitations associated with tractography in clinical practice. First, FGATIR allows high-resolution, isotropic, single-millimeter (3D) slice visualization of DBS targets with increased contrast-to-noise ratio.^45^ In a clinical setting, optimal dMRI currently features resolutions of 1.5 mm at best. Second, FGATIR is comparatively easy to implement as acquisition does not require specialized equipment, personnel or postprocessing techniques. FGATIR can be readily applied and is directly readable by any clinical PACS system or stereotactic planning software. The FGATIR protocol, which was employed in the present study can be obtained from the Supplementary Material. Third, although the acquisition of FGATIR is considerably longer than that of T1w and T2w sequences, it is still more time-efficient than conventional dMRI protocols, which may require 3-to 9-fold longer scanning times.^46^ Finally, identification of DRT by means of tractography yields major streamlines spanning from cerebellum to motor cortex (Fig 2D) that disregard synaptic relay stations and decussating fibers. Owing to a lack of anatomical constraints, specifically in the ventrodorsal plane, this impedes the identification of a distinct surgical target during planning. In contrast, FGATIR features a confined visual marker within PSA that has an approximate volume of 125 mm^3^ and is readily identifiable. This offers the potential for consistent targeting of the aforementioned hypointensity in all three planes. Indeed, our findings suggest that a trajectory along the major axis of the hypointensity might constitute the most optimal targeting strategy (Fig 4). This hypothesis, however, remains to be validated prospectively, especially with respect to potential side-effects associated with stimulation of internal capsule, lemniscal system, and red nucleus.

Another important consideration with respect to visualization of DRT is the bipartite organization of the outflow tract. Specifically, DRT features a decussating portion (dDRT) that makes up two-thirds of the ascending cerebellothalamic fibers and reaches the contralateral thalamus after decussation at the midbrain level.^23^ A smaller portion, however, does not decussate (ndDRT) and maintains an ipsilateral course that is distinct from dDRT.^26,28^ While the precise role of ndDRT in tremor pathophysiology remains to be established it is important to note that the tract frequently serves as a surrogate for DRT proper in dMRI-based targeting owing to the challenge of modeling the crossing fibers of dDRT (see also Fig 2D).^3,43^ These approaches have assumed a convergence of both tracts during their ascension into thalamus, as evidenced by recent tractography and microdissection studies^47^; the precise level where this intersection occurs, however, cannot be derived easily based on dMRI. As demonstrated in Figure 2F, our visual marker might coincide with the convergence of dDRT and ndDRT. Indeed, the strong hypointense signal appreciable on FGATIR sequences might exactly represent the approximation of both tracts as reflected by the elevated degree of myelination.^12^ Of note, the most efficacious hotspots derived from other clinical studies also clustered in this area, featuring distinct overlap with the identified hypointensity and likely encapsulating both dDRT and ndDRT during electrical stimulation (Fig 4). It is important to note, however, that in our sample, stimulating dDRT (R^2^= 0.03, p = 0.354) and ndDRT (R^2^= 0.001, p = 0.866) based on the anatomical definitions of the two tract atlases by Middlebrooks et al.^26^ were not able to significantly explain variance in clinical outcome. In summary, further investigation of the exact substrates underlying the observed hypointensity is needed. Diffusion MRI might constitute an optimal tool for this investigation, however, given that no dMRI data was available in our study population, future studies remain to investigate dMRI and FGATIR sequences in a single cohort. For now, we could establish that the spatial hypointensity marker is highly promising for clinical use with the potential to guide DBS programming and surgical targeting.

In recent years, an increasing emphasis has been placed on group-level analysis to identify the substrates associated with optimal stimulation outcome and to generate predictive models on a group level.^13,28^ However, these analyses have – by necessity – been conducted in MNI space. While advanced nonlinear registration pipelines are capable of incorporating *parts* of interindividual anatomical variability even in MNI space, a significant fraction of patient-specific information is usually lost. This holds especially true for complex multi-nucleus structures with low contrast-to-noise ratio, such as the thalamus. Here, co-registration often yields a blurry amalgamation of individual nuclei that lacks precision and anatomical detail. In consequence, despite having proven successful at explaining variance in clinical outcome in out-of-sample data, the translation of group-level findings into clinical practice remains a considerable challenge – especially in VIM-DBS. Advanced refinement tools – e.g. with manual refinements of warp fields^48^ – may be able to partially account for these registration errors and facilitate a smooth transition between MNI space and native space at some point. However, based on the findings of the present study, we advocate for the use of personalized DBS models in VIM DBS, especially if the aim is to translate findings into clinical practice. Specifically, our results indicate that transformation of native data into MNI space yields correlations strengths that are comparable to analysis in native space. In contrast, analysis based on atlas data and derived templates explained significantly less amount of variance (Fig. 3).

## Limitations

This study has several limitations pertaining to the interrogation and validation of advanced imaging sequences for DBS targeting. First, the data associated with the reported findings was gathered retrospectively and the clinical value and outcome of FGATIR-based VIM-targeting remains to be established in future prospective trials. This holds especially true for stimulation-induced side-effects, which were not directly investigated in the present study, but could potentially prevent targeting and stimulation of the identified hypointensity in some patients. Indeed, while studies have demonstrated exquisite tremor control within PSA, the low threshold for side effects such as dysarthria, dysmetria, ataxia, and paresthesia may reduce the therapeutic window of stimulation and limit the efficacy of this target compared to VIM proper. This issue could become more pertinent during chronic stimulation (typically > 3 years), where ET patients frequently experience waning stimulation benefit requiring larger currents and frequent reprogramming to account for disease progression and habituation to stimulation. The efficacy of PSA stimulation within the confines of the identified hypointensity under these circumstances has not been investigated in the present study and remains to be established. Second, to account for interindividual anatomical variability we investigated the predictive power of our identified hypointensity in native space. However, only a minority of patients (n=29) with sufficiently long follow-up could be included. Hence, future studies remain to establish normative vs. patient-specific differences in larger cohorts. Third, with an overall volume of 124.9 ± 35.2 mm^3^ the identified hypointensity features a considerable size and the optimal targeting strategy during surgical planning remains to be established. The results of our study, however suggest that volume overlap of electric fields with the identified hypointensity as well as distance to the center of gravity constitute reliable predictor for reliable tremor suppression. Taking into account the spatial distribution of our identified hotspot and coordinates previously reported in the literature it appears that a trajectory along the major axis of the hypointensity might constitute an optimal targeting strategy. However, this remains to be validated prospectively. Fourth, it is important to emphasize the technical limitations associated with our image-processing pipeline and stimulation volume modeling approach. Specifically, inaccuracies may arise during transformation of electrodes into common space. By employing a sophisticated preprocessing pipeline and employing advanced concepts such as multispectral normalization^14^, brain shift correction^14^, and subcortical refinement^15^ we sought to reduce potential sources of error and ensure the highest possible registration accuracy at the thalamic level. In addition to image-preprocessing it is important to emphasize the limitation of our stimulation volume modeling approach, which was used to approximate the shape and size of electric fields. The model employed here is based on the relationship between activations of axon cable models and E-field magnitudes as established by Åström et al.^49^ While this model used standard impedance values and space tissue segmentations to estimate the volume of activated tissue, it remains a simplification of the manner wherein electrical current interfaces with the brain. Nonetheless, recent comparative work has indicated the general similarity of its results to computationally more intensive pathway activation models – at least as a first-order approximation.^50^ This is corroborated by several recent publications that have used this method and were able to cross-predict significant amounts of variance in clinical improvement in out-of-sample data.^9^

## Conclusion

This study identified, investigated and validated a novel imaging-derived marker that holds promise to inform targeting in ET. Specifically, an oval-shaped hypointensity that could be reliably detected on FGATIR MRI sequences showed robust predictive and clinical utility to define an optimal stimulation site effective for tremor suppression in ET. This finding could mark an important step in applying advanced imaging sequences in neuromodulation that have so far remained underutilized due to a lack of demonstrated clinical benefit. In contrast to susceptibility-based techniques and dMRI, FGATIR sequences are easy to implement, time efficient, show less distortion artifacts and do not rely on additional hardware or postprocessing pipelines, making them suitable for direct and widespread implementation even in smaller DBS centers. Hence, we argue that advanced imaging sequences such as the one investigated here will constitute an important adjunct in the neuromodulation armamentarium with the potential to change and refine surgical decision making and improve clinical outcome in the course.

## Data Availability

The FGATIR protocol is supplied in the supplementary material. The data that support the findings of this study are available from the corresponding author.

## Acknowledgments

This work was supported by the German Research Foundation (Deutsche Forschungsgemeinschaft, TRR 295) and the BIH-Charité Clinician Scientist Program funded by the Charité-Universitätsmedizin Berlin and the Berlin Institute of Health (DKu and DKr).

## Author Contributions

CN, GHS, and AH contributed to the conception and design of the study; CN, DKr, BA, LG, DKu, KF, UR, AT, TP, TMH, EHM, AAK, and AH contributed to the acquisition and analysis of the data; CN, DKr, BA, LG, DKu, KF, UR, AT, TP, TMH, EHM, AAK, GHS, and AH contributed to drafting the text or preparing the figures.

## REFERENCES

1. Boutet A, Loh A, Chow CT, et al. A literature review of magnetic resonance imaging sequence advancements in visualizing functional neurosurgery targets. J Neurosurg. Published online March 26, 2021:1–14. doi:10.3171/2020.8.JNS201125

2. Dembek TA, Petry-Schmelzer JN, Reker P, et al. PSA and VIM DBS efficiency in essential tremor depends on distance to the dentatorubrothalamic tract. NeuroImage Clin. 2020;26:102235. doi:10.1016/j.nicl.2020.102235

3. Coenen VA, Sajonz B, Prokop T, et al. The dentato-rubro-thalamic tract as the potential common deep brain stimulation target for tremor of various origin: an observational case series. Acta Neurochir (Wien). 2020;162(5):1053–1066. doi:10.1007/s00701-020-04248-2

4. Blomstedt P, Sandvik U, Tisch S. Deep brain stimulation in the posterior subthalamic area in the treatment of essential tremor. Mov Disord. 2010;25(10):1350–1356. doi:10.1002/mds.22758

5. Nowacki A, Debove I, Rossi F, et al. Targeting the posterior subthalamic area for essential tremor: proposal for MRI-based anatomical landmarks. J Neurosurg. 2019;131(3):820–827. doi:10.3171/2018.4.JNS18373

6. Nölte IS, Gerigk L, Al-Zghloul M, Groden C, Kerl HU. Visualization of the internal globus pallidus: sequence and orientation for deep brain stimulation using a standard installation protocol at 3.0 Tesla. Acta Neurochir (Wien). 2012;154(3):481–494. doi:10.1007/s00701-011-1242-8

7. Zrinzo L, Hariz M, Hyam JA, Foltynie T, Limousin P. Letter to the Editor: A paradigm shift toward MRI-guided and MRI-verified DBS surgery. J Neurosurg. 2016;124(4):1135–1138. doi:10.3171/2015.9.JNS152061

8. Lozano AM, Lipsman N, Bergman H, et al. Deep brain stimulation: current challenges and future directions. Nat Rev Neurol. 2019;15(3):148–160. doi:10.1038/s41582-018-0128-2

9. Horn A, Fox MD. Opportunities of connectomic neuromodulation. Neuroimage. 2020;221:117180. doi:10.1016/j.neuroimage.2020.117180

10. Li J, Li Y, Gutierrez L, et al. Imaging the Centromedian Thalamic Nucleus Using Quantitative Susceptibility Mapping. Front Hum Neurosci. 2020;13. doi:10.3389/fnhum.2019.00447

11. Vassal F, Coste J, Derost P, et al. Direct stereotactic targeting of the ventrointermediate nucleus of the thalamus based on anatomic 1.5-T MRI mapping with a white matter attenuated inversion recovery (WAIR) sequence. Brain Stimul. 2012;5(4):625–633. doi:10.1016/j.brs.2011.10.007

12. Lehman VT, Lee KH, Klassen BT, et al. MRI and tractography techniques to localize the ventral intermediate nucleus and dentatorubrothalamic tract for deep brain stimulation and MR-guided focused ultrasound: A narrative review and update. Neurosurg Focus. 2020;49(1):E8. doi:10.3171/2020.4.FOCUS20170

13. Al-Fatly B, Ewert S, Kübler D, Kroneberg D, Horn A, Kühn AA. Connectivity profile of thalamic deep brain stimulation to effectively treat essential tremor. Brain. 2019;142(10):3086–3098. doi:10.1093/brain/awz236

14. Horn A, Li N, Dembek TA, et al. Lead-DBS v2: Towards a comprehensive pipeline for deep brain stimulation imaging. Neuroimage. Published online 2019. doi:10.1016/j.neuroimage.2018.08.068

15. Avants BB, Tustison NJ, Song G, Cook PA, Klein A, Gee JC. A reproducible evaluation of ANTs similarity metric performance in brain image registration. Neuroimage. 2011;54(3):2033–2044. doi:10.1016/j.neuroimage.2010.09.025

16. Vorwerk J, Oostenveld R, Piastra MC, Magyari L, Wolters CH. The FieldTrip-SimBio pipeline for EEG forward solutions. Biomed Eng Online. 2018;17(1):37. doi:10.1186/s12938-018-0463-y

17. Elias GJB, Boutet A, Joel SE, et al. Probabilistic Mapping of Deep Brain Stimulation: Insights from 15 Years of Therapy. Ann Neurol. Published online 2020. doi:10.1002/ana.25975

18. Su JH, Thomas FT, Kasoff WS, et al. Thalamus Optimized Multi Atlas Segmentation (THOMAS): fast, fully automated segmentation of thalamic nuclei from structural MRI. Neuroimage. 2019;194:272–282. doi:10.1016/j.neuroimage.2019.03.021

19. Horn A, Kühn AA, Merkl A, Shih L, Alterman R, Fox M. Probabilistic conversion of neurosurgical DBS electrode coordinates into MNI space. Neuroimage. 2017;150:395–404. doi:10.1016/j.neuroimage.2017.02.004

20. Ilinsky I, Horn A, Paul-Gilloteaux P, Gressens P, Verney C, Kultas-Ilinsky K. Human motor thalamus reconstructed in 3D from continuous sagittal sections with identified subcortical afferent territories. eNeuro. 2018;5(3):0060–18. doi:10.1523/ENEURO.0060-18.2018

21. Ewert S, Plettig P, Li N, et al. Toward defining deep brain stimulation targets in MNI space: A subcortical atlas based on multimodal MRI, histology and structural connectivity. Neuroimage. 2018;170:271–282. doi:10.1016/j.neuroimage.2017.05.015

22. Schaltenbrand G, Wahren W, Hassler R. Atlas for Stereotaxy of the Human Brain. 2nd ed. Thieme; 1977.

23. Neudorfer C, Maarouf M. Neuroanatomical background and functional considerations for stereotactic interventions in the H fields of Forel. Brain Struct Funct. 2018;223(1). doi:10.1007/s00429-017-1570-4

24. Sandvik U, Koskinen L-O, Lundquist A, Blomstedt P. Thalamic and subthalamic deep brain stimulation for essential tremor: where is the optimal target? Neurosurgery. 2012;70(4):840–845; discussion 845-6. doi:10.1227/NEU.0b013e318236a809

25. Petersen M V., Mlakar J, Haber SN, et al. Holographic Reconstruction of Axonal Pathways in the Human Brain. Neuron. 2019;104(6):1056-1064.e3. doi:10.1016/j.neuron.2019.09.030

26. Middlebrooks EH, Domingo RA, Vivas-Buitrago T, et al. Neuroimaging Advances in Deep Brain Stimulation: Review of Indications, Anatomy, and Brain Connectomics. Am J Neuroradiol. 2020;41(9):1558–1568. doi:10.3174/ajnr.A6693

27. García-Gomar MG, Soto-Abraham J, Velasco-Campos F, Concha L. Anatomic characterization of prelemniscal radiations by probabilistic tractography: implications in Parkinson’s disease. Brain Struct Funct. 2017;222(1):71–81. doi:10.1007/s00429-016-1201-5

28. Tsuboi T, Wong JK, Eisinger RS, et al. Comparative connectivity correlates of dystonic and essential tremor deep brain stimulation. Brain. Published online April 23, 2021. doi:10.1093/brain/awab074

29. Fytagoridis A, Åström M, Samuelsson J, Blomstedt P. Deep Brain Stimulation of the Caudal Zona Incerta: Tremor Control in Relation to the Location of Stimulation Fields. Stereotact Funct Neurosurg. 2016;94(6):363–370. doi:10.1159/000448926

30. Philipson J, Blomstedt P, Hariz M, Jahanshahi M. Deep brain stimulation in the caudal zona incerta in patients with essential tremor: effects on cognition 1 year after surgery. J Neurosurg. 2021;134(1):208–215. doi:10.3171/2019.9.JNS191646

31. Papavassiliou E, Rau G, Heath S, et al. Thalamic deep brain stimulation for essential tremor: relation of lead location to outcome. Neurosurgery. 2004;54(5):1120-1129; discussion 1129-30. doi:10.1227/01.neu.0000119329.66931.9e

32. Hamel W, Herzog J, Kopper F, et al. Deep brain stimulation in the subthalamic area is more effective than nucleus ventralis intermedius stimulation for bilateral intention tremor. Acta Neurochir (Wien). 2007;149(8):749–758. doi:10.1007/s00701-007-1230-1

33. Herzog J, Hamel W, Wenzelburger R, et al. Kinematic analysis of thalamic versus subthalamic neurostimulation in postural and intention tremor. Brain. 2007;130(6):1608–1625. doi:10.1093/brain/awm077

34. Barbe MT, Liebhart L, Runge M, et al. Deep brain stimulation of the ventral intermediate nucleus in patients with essential tremor: Stimulation below intercommissural line is more efficient but equally effective as stimulation above. Exp Neurol. 2011;230(1):131–137. doi:10.1016/j.expneurol.2011.04.005

35. Cury RG, Fraix V, Castrioto A, et al. Thalamic deep brain stimulation for tremor in Parkinson disease, essential tremor, and dystonia. Neurology. 2017;89(13):1416–1423. doi:10.1212/WNL.0000000000004295

36. Fiechter M, Nowacki A, Oertel MF, et al. Deep Brain Stimulation for Tremor: Is There a Common Structure? Stereotact Funct Neurosurg. 2017;95(4):243–250. doi:10.1159/000478270

37. Barbe MT, Reker P, Hamacher S, et al. DBS of the PSA and the VIM in essential tremor. Neurology. 2018;91(6):e543–e550. doi:10.1212/WNL.0000000000005956

38. Nowacki A, Bogdanovic M, Sarangmat N, Fitzgerald J, Green A, Aziz TZ. Revisiting the rules for anatomical targeting of ventralis intermediate nucleus. J Clin Neurosci. 2019;68:97–100. doi:10.1016/j.jocn.2019.07.027

39. Tsuboi T, Jabarkheel Z, Zeilman PR, et al. Longitudinal follow-up with VIM thalamic deep brain stimulation for dystonic or essential tremor. Neurology. 2020;94(10):e1073–e1084. doi:10.1212/WNL.0000000000008875

40. Middlebrooks EH, Okromelidze L, Wong JK, et al. Connectivity Correlates Predicting Deep Brain Stimulation Outcome in Essential Tremor: Evidence for a common treatment pathway. NeuroImage Clin. 2021;under Revi.

41. Marsden JF, Ashby P, Limousin-Dowsey P, Rothwell JC, Brown P. Coherence between cerebellar thalamus, cortex and muscle in man: Cerebellar thalamus interactions. Brain. 2000;123(7):1459–1470. doi:10.1093/brain/123.7.1459

42. Petry-Schmelzer JN, Dembek TA, Steffen JK, et al. Selecting the Most Effective DBS Contact in Essential Tremor Patients Based on Individual Tractography. Brain Sci. 2020;10(12):1015. doi:10.3390/brainsci10121015

43. Sammartino F, Krishna V, King NKK, et al. Tractography-Based Ventral Intermediate Nucleus Targeting: Novel Methodology and Intraoperative Validation. Mov Disord. 2016;31(8):1217–1225. doi:10.1002/mds.26633

44. Behrens TEJ, Berg HJ, Jbabdi S, Rushworth MFS, Woolrich MW. Probabilistic diffusion tractography with multiple fibre orientations: What can we gain? Neuroimage. 2007;34(1):144–155. doi:10.1016/j.neuroimage.2006.09.018

45. Sudhyadhom A, Haq IU, Foote KD, Okun MS, Bova FJ. A high resolution and high contrast MRI for differentiation of subcortical structures for DBS targeting: The Fast Gray Matter Acquisition T1 Inversion Recovery (FGATIR). Neuroimage. 2009;47(2):44–52. doi:10.1016/j.neuroimage.2009.04.018

46. Petersen M V., Lund TE, Sunde N, et al. Probabilistic versus deterministic tractography for delineation of the cortico-subthalamic hyperdirect pathway in patients with Parkinson disease selected for deep brain stimulation. J Neurosurg. 2017;126(5):1657–1668. doi:10.3171/2016.4.JNS1624

47. Petersen KJ, Reid JA, Chakravorti S, et al. Structural and functional connectivity of the nondecussating dentato-rubro-thalamic tract. Neuroimage. 2018;176:364–371. doi:10.1016/j.neuroimage.2018.04.074

48. Edlow B, Mareyam A, Horn A, et al. 7 Tesla MRI of the ex vivo human brain at 100 micron resolution. Sci Data. 2019;6(244). doi:10.1101/649822

49. Åström M, Diczfalusy E, Martens H, WÅrdell K. Relationship between neural activation and electric field distribution during deep brain stimulation. IEEE Trans Biomed Eng. 2015;62(2):664–672. doi:10.1109/TBME.2014.2363494

50. Duffley G, Anderson DN, Vorwerk J, Dorval AD, Butson CR. Evaluation of methodologies for computing the deep brain stimulation volume of tissue activated. J Neural Eng. 2019;16(6):066024. doi:10.1088/1741-2552/ab3c95

