## Supplementary Material for "Personalizing deep brain stimulation using advanced imaging sequences"

### SIEMENS MAGNETOM Skyra

#### Routine

| | |
|---|---|
| Slab group 1 | |
| Slabs | 1 |
| Dist. factor | 50 [%] |
| Position | Isocenter |
| Orientation | P6.8 P6.1 H7.2 mm |
| Phase enc. dir. | R >> L |
| Rotation | 90 [deg] |
| Phase oversampling | 0 [%] |
| Slice oversampling | 0 [%] |
| Slices per slab | 160 |
| FoV read | 240 [mm] |
| FoV phase | 81.3 [%] |
| Slice thickness | 1 [mm] |
| TR | 3000 [ms] |
| TE | 3.44 [ms] |
| Averages | 1 |
| Concatenations | 1 |
| Filter | Image filter, ... |
| Coil elements | HE1-4 |

#### Contrast

| | |
|---|---|
| Magn. preparation | Non-sel. IR |
| TI | 414 [ms] |
| Flip angle | 8 [deg] |
| Reconstruction | Magnitude |
| Fat suppr. | None |
| Water suppr. | None |
| Measurements | 1 |

#### Resolution

| | |
|---|---|
| Base resolution | 256 |
| Phase resolution | 100 [%] |
| Slice resolution | 100 [%] |
| Phase partial Fourier | 7/8 |
| Slice partial Fourier | 6/8 |
| Filter 1 | |
| Raw filter | Off |
| Filter 2 | |
| Large FoV | On |
| Filter 3 | |
| Normalize | Off |
| Filter 4 | |
| Elliptical filter | Off |
| Interpolation | Off |
| PAT mode | None |

#### Geometry

| | |
|---|---|
| Multi-slice mode | Single shot |
| Series | Interleaved |

#### System

| | |
|---|---|
| Save uncombined | 0 |
| Scan at current TP | 0 |
| Scan region position | H |
| Scan region position | 0 [mm] |
| MSMA | S - C - T |
| Sagittal | R >> L |
| Coronal | A >> P |
| Transversal | F >> H |
| Head 3T / HE | 1 |
| Shim mode | Standard |
| Confirm freq. adjustment | 0 |
| Assume Silicone | 0 |
| Ref. amplitude [1H] | 126.364 [V] |
| Adjust volume | |
| Position | Isocenter |
| Orientation | Transversal |
| Rotation | 0.0[deg] |
| A >> P | 263 [mm] |
| R >> L | 350 [mm] |
| F >> H | 350 [mm] |

#### Physio

| | |
|---|---|
| 1st Signal/Mode | None |
| Dark blood | 0 |
| Resp. control | Off |

#### Inline

| | |
|---|---|
| Subtract | 0 |
| Std-Dev-Sag | 0 |
| Std-Dev-Cor | 0 |
| Std-Dev-Tra | 0 |
| Std-Dev-Time | 0 |
| MIP-Sag | 0 |
| MIP-Cor | 0 |
| MIP-Tra | 0 |
| MIP-Time | 0 |
| Save original images | 1 |

#### Sequence

| | |
|---|---|
| Introduction | 1 |
| Dimension | 3D |
| Elliptical scanning | 0 |
| Averaging mode | Long term |
| Asymmetric echo | Allowed |
| Bandwidth | 130 [Hz/Px] |
| Echo spacing | 9.7 [ms] |
| RF pulse type | Fast |
| Gradient mode | Fast |
| Excitation | Slab-sel. |
| RF spoiling | 1 |
